# Indoor air quality at a French university: *a participatory CO2 measurement campaign highlights the wide gap between reality and the law*

**DOI:** 10.1101/2025.01.28.25321193

**Authors:** Philippe Cinquin

**Affiliations:** Univ. Grenoble Alpes, CNRS, UMR 5525, VetAgro Sup, Grenoble INP, CHU Grenoble Alpes, TIMC, UMR5525, 38000 Grenoble, France

## Abstract

Poor indoor air quality has been demonstrated to increase the risk of transmitting infectious agents and to expose individuals to the phenomenon of sick building syndrome. In light of these findings, most countries have established specific guidelines regarding indoor air quality in university rooms. In France, for instance, the maximum permissible concentration of carbon dioxide (CO_2_) in university rooms without mechanical ventilation is set at 1,300 parts per million (ppm), and the minimum volume per occupant is 15 m^3^. For rooms with mechanical ventilation, the minimum clean air flow rate is 25 m^3^/h/occupant.

The primary objective of this study was to design and demonstrate the feasibility of a simple, cost-effective method for comparing the reality of indoor air quality in all university rooms with legal requirements. The secondary objectives of the present study were to demonstrate the efficacy of the proposed method in identifying and reporting problematic situations, and in issuing practical recommendations.

Mobile CO_2_ sensors (Aranet4) were provided to volunteer lecturers to measure the CO_2_ concentration during and after classes. The number of occupants and the condition of openings were also recorded. These data were supplemented by measurements from 117 fixed Carbon Nexelec sensors. The data were then fitted to a model, which enabled the characterization of air quality and the estimation of the gauge reduction required to comply with the law.

None of the 14 rooms without mechanical ventilation complied with the legal minimum of 15 m^3^/occupant. 75% of the third quartiles of CO_2_ concentrations during classes exceeded 2692 ppm. In rooms with mechanical ventilation, median clean air flow was 15 m^3^/h/occupant at 100% occupancy (9 m^3^/h/occupant for the first quartile). In 32 out of 41 rooms with mechanical ventilation (78%), the clean air flow was estimated to be below the legal minimum of 25 m^3^/h/occupant at 100% occupancy. Concentrations in excess of 5,000 ppm were observed in 23 of the 101 rooms equipped with fixed sensors.

The proposed method has demonstrated its feasibility in real-life conditions. For the purpose of evaluating the air quality of all rooms affiliated with universities, it is recommended that this method be used in a systematic manner. The findings of this study indicate that a significant proportion of the examined rooms may not be in accordance with the relevant legislation, thereby jeopardizing the health of the occupants. In order to comply with the law, the method proposed here to estimate gauge reduction should be applied.

## 1. Introduction

The Corona Virus Disease 2019 (CoViD-19) pandemic has drawn attention to the risk of airborne transmission of infectious agents, particularly in enclosed environments. Indeed, as early as 2020, several studies highlighted the airborne transmission of the severe acute respiratory syndrome coronavirus 2, SARS-CoV-2 [1] [2]. It therefore immediately became clear that the quality of ventilation in the workplace was an essential factor in limiting the spread of the pandemic. Indeed, several studies had confirmed that the incidence rate of airborne infectious diseases is higher in poorly ventilated premises than in well-ventilated premises [3] [4]. Since then, studies have confirmed that the installation of effective mechanical ventilation in school classrooms significantly reduces the risk of SARS-CoV-2 contamination [5].

However, it has long been recognized that carbon dioxide (CO_2_) concentration can be used to assess the quality of ventilation in a room [6]. More specifically, a link has been established between a high CO_2_ concentration and an increased risk of transmission of airborne viruses such as SARS-CoV-2 [7]. Furthermore, physico-chemical mechanisms explaining the effect on SARS-CoV-2 infectivity of even a moderate increase in CO_2_ concentration are beginning to be elucidated [8].

Moreover, independently of the increased risk of transmission of airborne infections, various studies report a correlation between high CO_2_ concentrations and various psycho-cognitive disorders, including headaches, fatigue, nausea and even dizziness. Sick Building Syndrome [9], a nosological entity recognized by the World Health Organization since 1982, brings together the various symptoms experienced by occupants of poorly ventilated buildings. These symptoms may be linked to CO_2_ itself or to various chemical pollutants present in the atmosphere of public buildings, particularly Volatile Organic Compounds exhaled by building occupants [10], [11], [12].

For all these reasons, legislation in many countries sets maximum CO_2_ concentration thresholds not to be exceeded, as well as minimum ventilation rates in rooms with mechanical ventilation, compatible with the recommendations of the American Society of Heating, Refrigerating and Air-Conditioning Engineers [6]. This is the case in France, where the following thresholds apply in universities. In university rooms with mechanical ventilation, the legal maximum CO_2_ concentration is 1,000 parts per million (ppm), with a tolerance of up to 1,300 ppm, and the minimum clean air flow per occupant is 25 m^3^/h/occupant. In university rooms without mechanical ventilation, there is no legal maximum CO_2_ concentration, however a minimum volume per occupant of 15 m^3^/occupant is required. In two opinions [13] [14], France’s highest public health authority, the HCSP (High Council for Public Health), has set a “rapid action threshold” of 1,500 ppm “*indicating unacceptable air confinement in the light of the scientific literature, and requiring corrective action (lowering the occupancy gauge or evacuating the room, modifying the technical means of aeration and ventilation)*”. In a recent opinion [15], it set a maximum CO_2_ concentration target of 800 ppm and a minimum clean air flow per occupant in mechanically ventilated rooms of 50 m^3^/h/occupant.

The public health benefits of working in good-quality air have therefore been demonstrated, and the legal framework guaranteeing air quality in buildings open to the public, particularly universities, is clear. However, to the best of our knowledge, no study has proposed a method for measuring indoor air quality in universities that is designed to check the compatibility of all university classrooms with the relevant legal requirements. A number of one-off studies have been carried out to characterize CO_2_ concentrations or fresh-air ventilation rates in selected university or school classrooms (see, for example, [3] [4] [5] [16] [17]). [17] delineates three classes of methods applicable to estimating fresh air ventilation rates: direct measurement of flow, study of the decay of marker gases diffused in a controlled manner, and study of markers **-**in particular CO_2_**-** spontaneously emitted when rooms are used by humans. [17] characterized the ventilation of 214 rooms in seven universities or schools, most of which had mechanical ventilation. However, the majority of the estimates of ventilation performance in this study were derived from the study of the decay of marker gases, which would be incompatible with the systematic and regular estimation of all ventilation rates.

The primary objective of the present study was to demonstrate the feasibility of a method designed to ascertain the compatibility of all university rooms with legal requirements in terms of CO_2_ concentration and fresh air ventilation rate. The implementation of this method on a broad scale would facilitate the determination of the CO_2_ concentration levels attained when university rooms are utilized under real-life conditions, as well as the estimation of the performance of both natural and mechanical ventilation systems. The proposed method is sufficiently simple and inexpensive to be implemented on a very regular basis (at least once a year) in all rooms with mechanical ventilation. The air quality in these rooms is contingent on the performance of the air handling units, which exhibits variability over time due to the effectiveness of the maintenance procedures employed. This necessitates regular verification to ensure optimal functioning.

In order to achieve this objective, the present study applied a methodological approach to the analysis of the compatibility between the actual conditions of use of rooms in a French university, Univ. Grenoble Alpes (UGA), and the legal requirements and recommendations of the HCSP in terms of CO_2_ levels and ventilation performance. This entailed the characterization of the distributions of CO_2_ concentrations in all the rooms under study, contingent on their actual conditions of use, as well as the mechanical ventilation performance of the rooms that had it or the volume per occupant in rooms lacking mechanical ventilation. A secondary objective was to demonstrate that our method enables detection of situations likely to endanger the health of users and alert the administrators of the universities. The third objective was to demonstrate the feasibility of using the proposed method to provide practical recommendations to university administrators regarding indoor air quality.

It swiftly became evident that to accomplish our objectives, it was imperative to collect not only CO_2_ values, but also data regarding actual room occupancy conditions (particularly the number of occupants and the state of door and window openings). These data are not readily available in the information systems of universities. While room reservation systems at universities facilitate the identification of rooms allocated for specific classes, the accuracy of predicting student attendance remains a challenge. While a network of fixed CO_2_ sensors offers valuable insights, it is not sufficient for the study’s objectives.

The collection of information necessary to achieve our objectives necessitates the commitment of teaching staff. This necessity led to the formulation of the original concept of a “participatory CO_2_ measurement campaign”. This concept is predicated on the principle of involving UGA staff in measuring CO_2_ (using portable sensors) and collecting data on actual classroom occupancy conditions.

## 2. Materials and Methods

### 2.1 Launch of a “participatory CO_2_ measurement campaign”

On October 23, 2023, the SNESUP-FSU union used its electronic mailing list for UGA staff to invite them to take part in a campaign to study CO_2_ under real conditions in UGA rooms. The pdf document to which they were directed presented the interest and objectives of the study, the commitments to be made in order to participate, how the results would be shared, and described the information to be gathered on the measurement conditions, as well as the method for implementing the sensor and sharing the measurements. Each participant had access to a personal section of the UGA cloud, where they could upload their CO_2_ measurement files for each session, as well as the file describing the measurement conditions. A reservation file accessible to all participants made it possible to reserve a sensor and to contact the colleague who had reserved it before and the colleague who had reserved it afterwards (so that participants could transmit sensors to each other, without centralized intervention). In the final part of the study, those who could were asked to leave the sensor in place for at least an hour after the end of the human presence in the room, with all openings closed.

### 2.2 Collecting room usage conditions as part of the “participatory campaign”

For each measurement session, each participant was asked to record the conditions of use of the room in a pre-determined format set out in an Excel template. This file asked for the following information: name of building, name of room, room gauge *J*, existence of mechanical ventilation, position of sensor, date, start time, end time, number of people in the room (as a function of time), state of opening of doors and windows (as a function of time), and if possible, room dimensions. Free-format observations could be added. For some rooms, measurements of room dimensions were taken using a Dexter laser rangefinder (accuracy ± 2 mm), enabling the area *A* and volume *V* of the room to be estimated.

### 2.3 CO_2_ measurements

#### 2.3.1 CO_2_ measurements using the “participatory campaign”

Four Aranet4 sensors were made available to participants. These sensors complied with the recommendations of the French Environment Code [18] for CO_2_ measurements (measurement by non-dispersive infrared absorption spectrometry, measurement range 0 to 5,000 ppm; measurement uncertainty ± [30 ppm + 3% of reading]). They measure CO_2_ concentrations in ppm. They had been calibrated in accordance with the manufacturer’s recommended procedure. This procedure consists of installing the sensor in a place where the CO_2_ concentration is close to 420 ppm. In order to be as close as possible to this value, the calibration was performed outdoors in an environment with adequate wind exposure, sparsely built-up, far from industrial activities and car traffic, with the sensor positioned at a distance from plants. The procedure is initiated from the application installed on a mobile phone and requires no intervention other than starting the calibration and waiting for it to complete (which takes about 15 minutes). Regular calibration checks were carried out, verifying that at certain times of the day they measured outdoor concentrations between 420 and 470 ppm. No recalibration was necessary. These lightweight, portable sensors record one measurement per minute, and digitally transfer the data in .csv format to a smartphone via an app that can be downloaded from any app store. Spot measurements were taken outside teaching buildings by some participants.

#### 2.3.2 Access to data collected by fixed UGA sensors

CO_2_ measurements from the “participatory campaign” were enriched by data captured by 117 Carbon Nexelec sensors deployed by the UGA. These sensors take a reading every 10 minutes and transmit their data over a private wireless long-range wide area network. The UGA makes all these measurements available to its staff, via its intranet, on a Grafana® platform.

### 2.4 Transfer of measurement conditions and measurements

The app downloaded by each participant onto their smartphone enabled them to transfer the measurement file to their personal computer via the application of their choice. For each measurement session, the participant uploaded two files to the personal section of the UGA cloud assigned to him/her: the CO_2_ measurement file, in .csv format; and the Excel file containing the measurement conditions.

### 2.5 Processing data from the “participatory campaign”

#### 2.5.1 Processing data measured during room occupancy

For each measurement session, an Excel interpretation file was created, containing both the measurement conditions and the measurements themselves. A graph of the CO_2_ curve versus time was generated, also showing the HCSP target values (800 and 1,500 ppm), the legal value (1,300 ppm), and the thresholds of 2,500 and 4,000 ppm. The following parameters were systematically calculated for each session, when the necessary information was available: gauge *J*, median *N* of the number of occupants, occupancy rate *R_Occ_* = *N/J*, volume per occupant when *R_Occ_* = 100% *V_J_ = V/J*, median of CO_2_, *Q_3_* value of the third quartile of CO_2_, portion of the duration of the session spent with CO_2_ > 1,500 ppm and with CO_2_ > 2,500 ppm, clean air flow per occupant *D_N_* when *N* occupants are present in the room (so *D_J_* is the clean air flow per occupant when *R_Occ_* = 100%), total clean air flow *D* in the room.

In order to estimate *D_N_*, the classical model of CO_2_ concentration in a well-mixed space was utilized [19]. This model states that, in a room with *N* occupants,

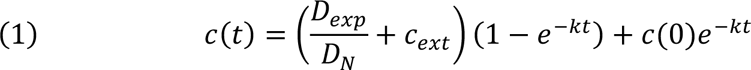

where *c(t)* is the concentration of CO_2_ at time *t*, *D_exp_* is the average CO_2_ generation rate by human beings, *c_ext_* is the concentration of CO_2_ outside the building, and 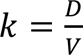 is the air renewal rate. As a consequence, when *c(t)* reaches a steady state *c_limit_*

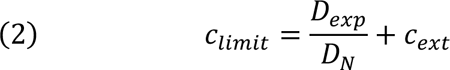

hence

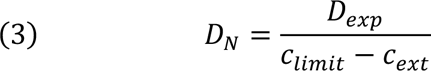

This model requires an estimation of *D_exp_*, the average CO_2_ generation rate by human beings. This rate has been determined to be 20.2 L/h [20]. We propose to estimate *c_limit_* by *Q_3_*. The reasons for these choices of the estimations of *D_exp_* and of *Q_3_* are discussed in section 4.1.2. As a consequence, if *c_ext_* is expressed in ppm, *D_N_* (m^3^/h) was estimated by

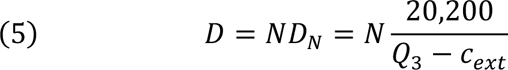

As a consequence,

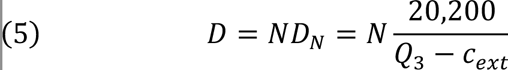

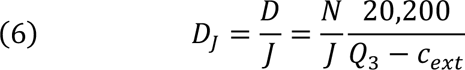

For university rooms without mechanical ventilation, French law imposes *V_J_ ≥ 15* m^3^/occupant. Accordingly, the legal gauge can be defined as *J_Legal_* = *min [J ; J V_J_ / 15]*. Although there is no legal maximal value of CO_2_ in university rooms in France, the HCSP Gauge *J_HCSP_* can be defined as the maximal number of occupants not to overpass the 1,500 ppm threshold recommended by HCSP. The previously mentioned model of CO_2_ concentration in a well-mixed space implies that, if the CO_2_ limit concentration is 1,500 ppm, the minimum flow of clean air per occupant *D_HCSP_* satisfies *D_HCSP_ = 20,200/(1,500-c_ext_)*, where *c_ext_* is the outdoor concentration. From this it can be deduced that *J_HCSP_ = min [J ; J D_J_ / D_HCSP_]*.

Since the true value of the outdoor CO_2_ concentration was rarely available, we opted to overestimate *D* by using the maximum value of CO_2_ measured outdoors (500 ppm), and to underestimate *D_HCSP_* by using the minimum possible value (417 ppm, world average CO_2_ concentration [21]). These choices all maximize the estimate of the *J_HCSP_* gauge. The reasons for this choice will be discussed in section 4.1.2.

For rooms with mechanical ventilation, French law imposes *D_J_ ≥* 25 m^3^/occupant in universities. The legal gauge can then be defined as *J_Legal_* = *min [J ; J D_J_ / 25]*.

#### 2.5.2 Processing data measured after room occupancy, with openings closed

The participant had to leave the CO_2_ sensor in the room for at least 1 hour, after the room had been used and CO_2_ had risen above the concentration outside the building. Equation (1) can be written as *c*(*t*) = *ae*^−*kt*^ + *c_ext_*. When *c_ext_* had been measured, this value was used. Otherwise, *c_ext_* was assumed to be 500 ppm (this leads to a limited overestimation of *k*, that will be discussed in section 4.1.2). To estimate parameters *a* and *k*, Excel’s SLOPE and INTERCEPT functions were used, on data {*t_i_*; *ln*(*c*(*_t_*) − *c_ext_*)}. Quality of fit was estimated by the correlation coefficient between measurements and model predictions, obtained by Excel’s CORREL function. Time being expressed in minutes, when the volume *V* of the room and its gauge *J* were known, the clean air flow per occupant at 100% occupancy was obtained by the formula *D_J_=60 kV/J*. The legal gauge *J_Legal_* is here also estimated by *J_Legal_* = *min [J ; J D_J_ /25]*.

### 2.6 Comparison of the results of the participatory campaign with data from sensors deployed by the UGA

In order to corroborate the results obtained by the participatory campaign, the measurements taken by the UGA sensors from September 2022 to June 2023 were utilized. The study was conducted in rooms where at least one measurement exceeded 5,000 ppm. Grafana® software was used to represent the CO_2_ concentration in these rooms over this period. From March 26 to April 16, 2024, the investigation’s primary focus was on analyzing rooms with CO_2_ levels exceeding 5,000 ppm. For these cases, Grafana’s .csv export function was employed, and the corresponding data were then exported to Excel. It was hypothesized that the human presence in these rooms would be characterized by a CO₂ concentration exceeding 550 ppm. For each of these rooms, a summary plot was produced, displaying the CO₂ concentrations above 550 ppm as a function of time. It was hypothesized that a quasi-linear decrease of log (*C_measured_* − *C_ext_*) over more than an hour after a CO₂ peak would indicate that the room was empty. To this end, the aforementioned method was employed to estimate the clean air renewal rate and the clean air flow per occupant at 100% occupancy. It is noteworthy that UGA’s Carbon Next sensors have an upper measurement limit of 5,000 ppm. The utilization of the model was undertaken for the purpose of predicting the maximum CO₂ value in the room during the “plateau” measured by the sensor. To this end, if time *t = 0* corresponds to the start of decay, the model *C*(*t*) = *ae*^−*kt*^ + *C_ext_* was employed with negative *t* times. The value of time corresponding to the actual end of room use was estimated using UGA’s room reservation management software. In this situation, it is interesting to plot not only the mean value of the model, but also its 95% confidence interval. The 95% confidence intervals for coefficients *a* and *k* were estimated using Excel’s “Data Analysis / Linear Regression” function. The production of two curves—low and high predictions—allowed the delineation of the 95% confidence interval for the predictions.

## 3 Results

### 3.1 Involvement in the participatory campaign

The participatory campaign involved 20 UGA staff, mainly from October 19, 2023 to December 21, 2023, with some additional measures in 2024. Three “participatory campaign letters” were distributed to participants, on November 1 and December 8, 2023, and February 21, 2024. CO_2_ was measured in 77 room-use sessions, in a total of 56 rooms in 21 buildings.

### 3.2 Conditions of the measurements of the participatory campaign

Most of the measurements were taken during a relatively cold period (midday temperature: October = 16°C; November = 6°C; December = 7°C). As a consequence, the heating was working, and the occupants were rather reluctant to open the windows. None of the rooms studied had air conditioning. The CO_2_ sensor was switched on by the teacher just before the course (by insertion of the batteries in the device). It was systematically placed on the teacher’s desk, at a distance of at least one meter from the teacher. At the end of the course, the teacher uploaded the CO_2_ measurement on his or her mobile phone, then on his or her personal section of the UGA cloud. Then the CO_2_ sensor was switched off by removing batteries.

### 3.3 Compliance with legal requirements and HCSP recommendations in rooms without mechanical ventilation surveyed by the participatory campaign

CO_2_ concentrations were studied in 20 classroom sessions in 15 rooms without mechanical ventilations, in 9 buildings of UGA. The volume was known for 14 of these 15 rooms. Table 1 summarizes the data recorded to estimate *J_Legal_* (legal gauge to meet the legal minimum of 15 m^3^/occupant). As can be seen from Table 1, none of these 14 rooms complied with the legal minimum of 15 m^3^/occupant.

**Table 1.**
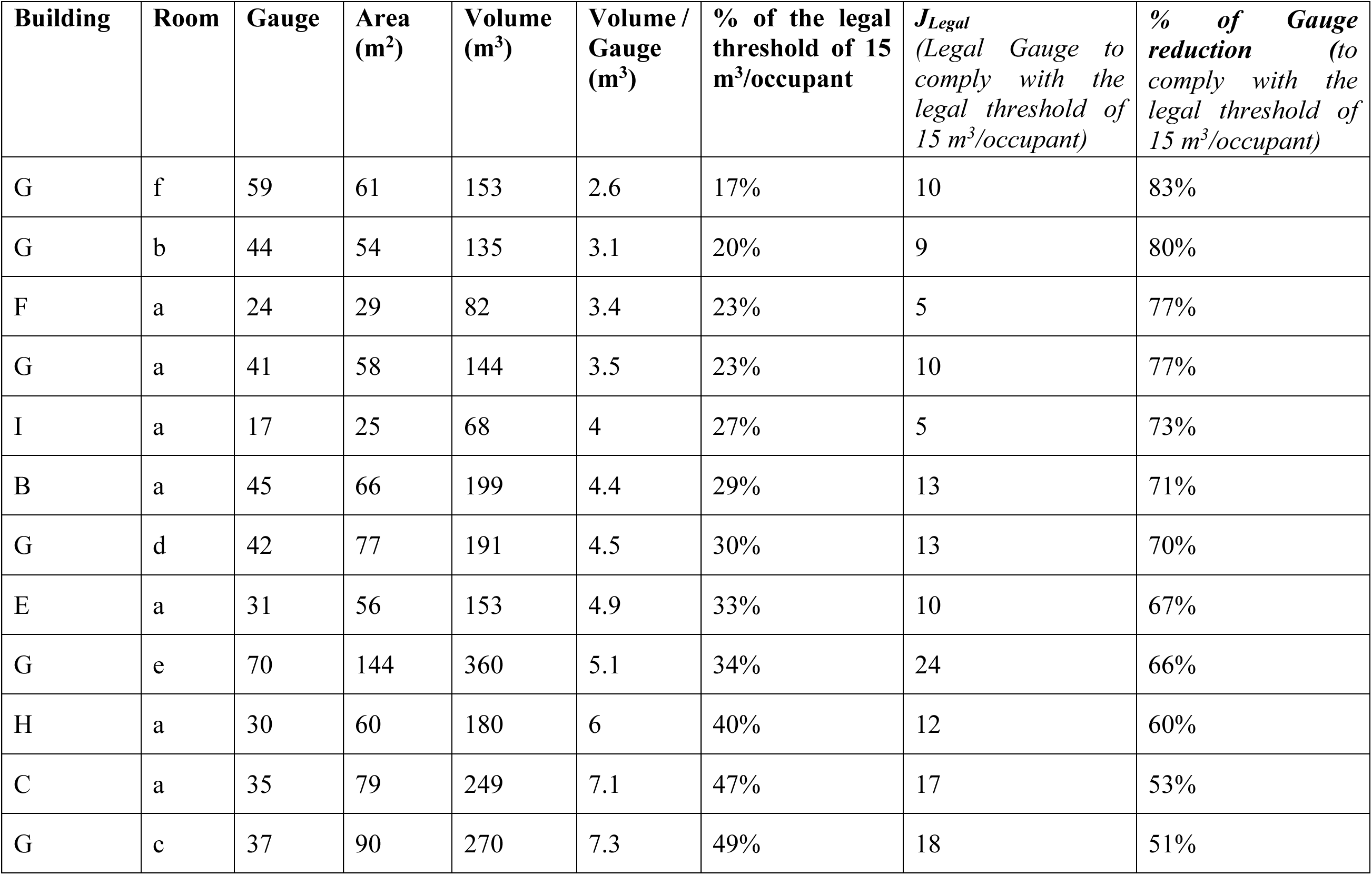
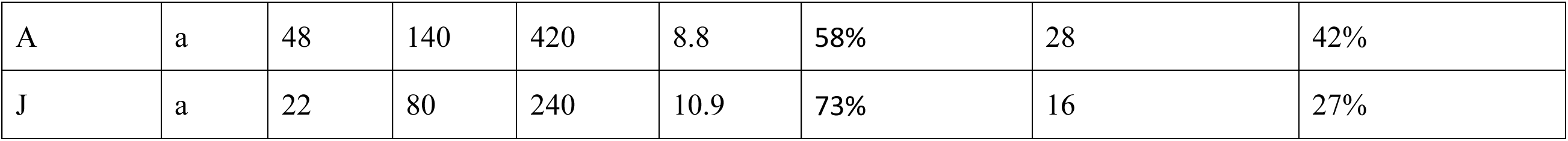
Summary of volume per occupant for the 14 rooms without mechanical ventilation for which the volume was known (sorted by the Volume / Gauge ratio), enabling estimation of the legal gauge required to respect the legal threshold of 15 m^3^/occupant.

Table SM.1 from the supplementary material summarizes the CO_2_ measurements made during the participatory campaign in the 15 rooms without mechanical ventilation, and the estimations of *J_HCSP_* (recommended gauge not to overpass the 1,500 ppm threshold recommended by HCSP).

The main parameters characterizing CO_2_ distribution in these rooms are synthesized in Table 2. In particular, 75% of the third quartiles of CO_2_ concentrations measured during room occupancy sessions were above 2,692 ppm.

**Table 2.**
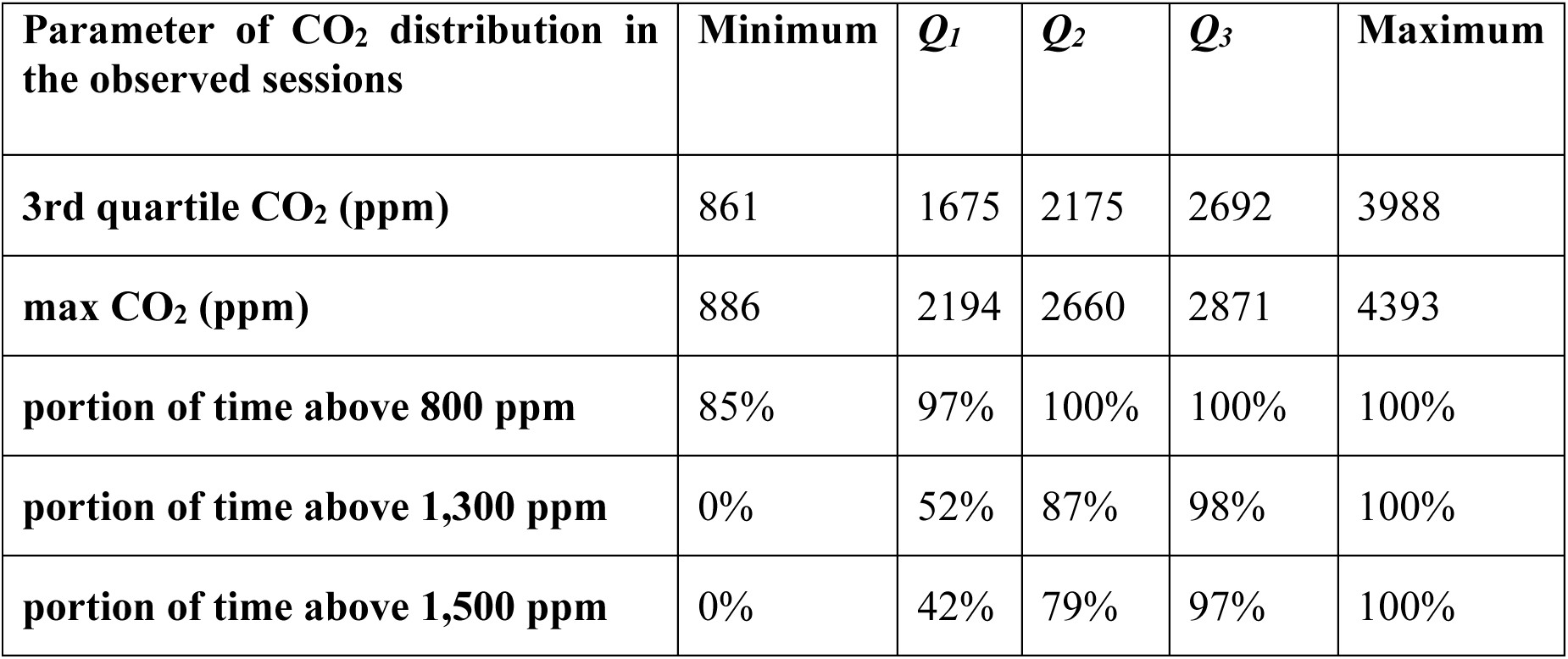
Synthetic description of the main parameters characterizing the distribution of CO_2_ measurements in the 15 rooms without mechanical ventilation.

| Parameter of CO<sub>2</sub> distribution in the observed sessions | Minimum | $Q_1$ | $Q_2$ | $Q_3$ | Maximum |
| --- | --- | --- | --- | --- | --- |
| 3rd quartile CO<sub>2</sub> (ppm) | 861 | 1675 | 2175 | 2692 | 3988 |
| max CO<sub>2</sub> (ppm) | 886 | 2194 | 2660 | 2871 | 4393 |
| portion of time above 800 ppm | 85% | 97% | 100% | 100% | 100% |
| portion of time above 1,300 ppm | 0% | 52% | 87% | 98% | 100% |
| portion of time above 1,500 ppm | 0% | 42% | 79% | 97% | 100% |

Typical examples of the effect of ventilation are shown in Figures 1 and 2.

**Figure 1.**
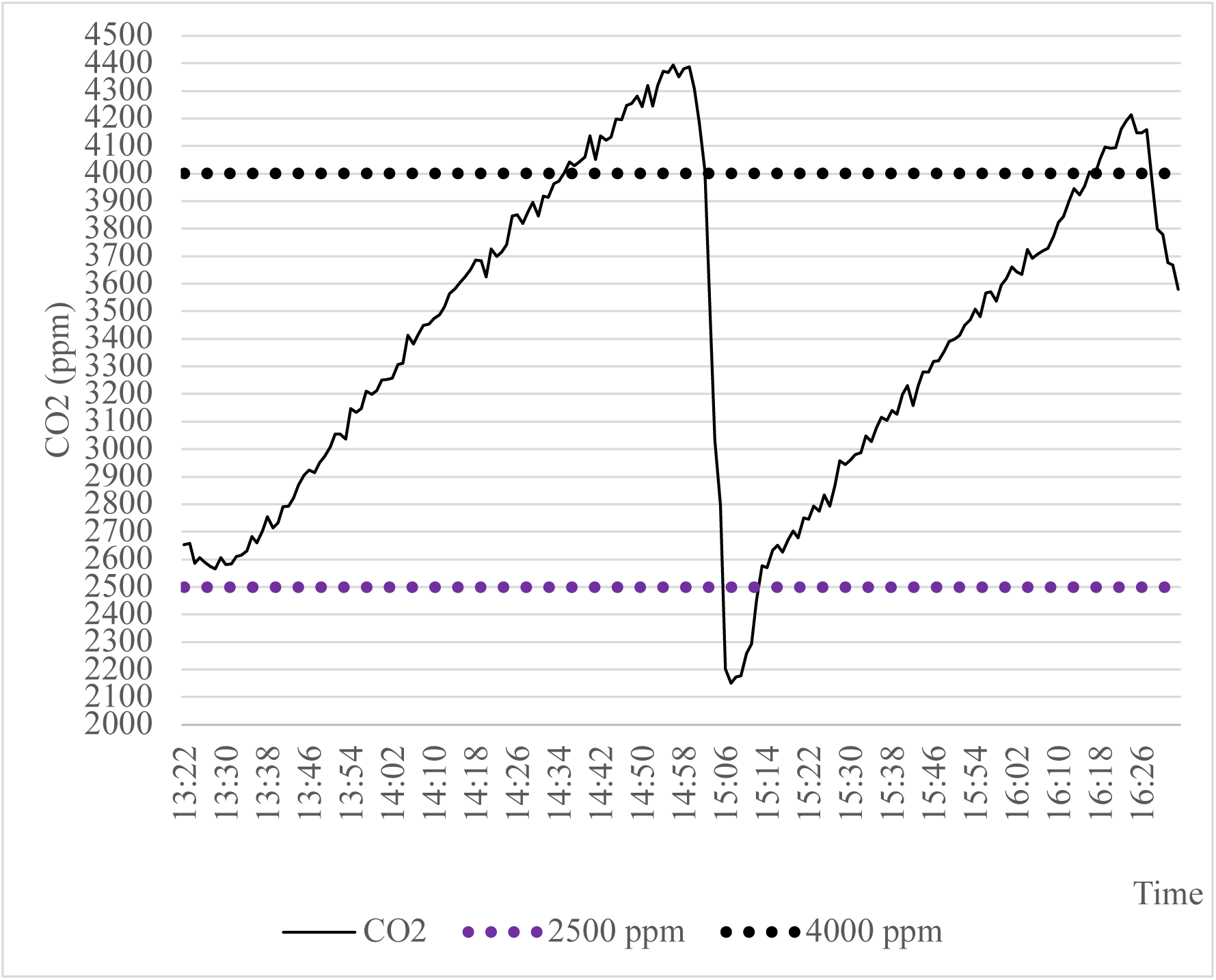
Typical example of CO_2_ growth in room Aa, a room of 420 m^3^ (14 m x 10 m x 3 m), with a gauge of 48, without mechanical ventilation. All three doors and 5 windows were closed during the two sessions. 21 students and one teacher were present (19 m^3^/occupant, 46% of gauge). During the 15 minutes break, one door and two windows were opened, and 11 students left the room.

**Figure 2.**
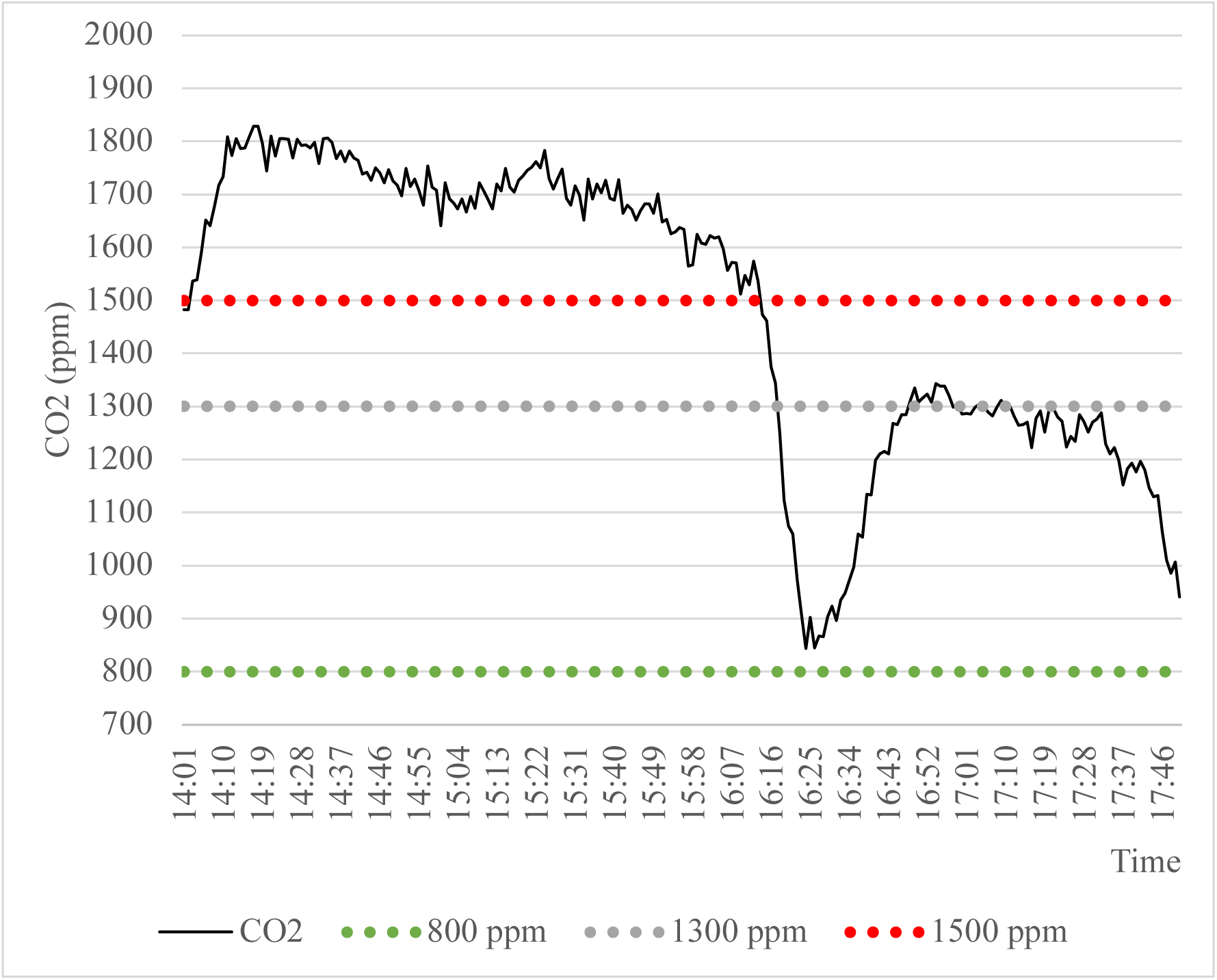
Illustration of the importance of a draught, even when windows are open. Room Ia is 67.5 m^3^ (5 m x 5 m x 2.7 m), with a gauge of 17. 20 persons were present (118 % of the gauge). The volume per occupant is 3.4 m^3^/occupant, which is 23% of the legal minimum of 15 m^3^/occupant. The only door is closed during the sessions, and the 4 windows are open. The opening of the windows keeps CO_2_ at a plateau (which is however above the HCSP recommendation of 1,500 ppm). During the break, the installation of a draught by opening of the door significantly reduces the CO_2_ concentration.

### 3.4 Compliance with legal requirements and HCSP recommendations in the mechanically ventilated rooms surveyed by the participatory campaign

#### 3.4.1 Study of data collected during room use

CO_2_ concentrations were studied during 57 classroom sessions in 41 mechanically ventilated rooms in 15 UGA buildings. The main parameters describing characteristics and room occupancy, CO_2_ distribution and mechanical ventilation performance in these rooms are synthesized in Table 3.

**Table 3.** Synthetic description of room characteristics and occupancy, CO_2_ distribution and mechanical ventilation, in 57 classroom sessions in 41 rooms with mechanical ventilation, in 15 UGA buildings.

| | Minimum | $Q_1$ | $Q_2$ | $Q_3$ | Maximum |
| --- | --- | --- | --- | --- | --- |
| **Characteristics of the measurement sessions** |  |  |  |  |  |
| Room volume (m<sup>3</sup>) | 62 | 126 | 178 | 900 | 1360 |
| Room area (m<sup>2</sup>) | 21 | 50 | 70 | 215 | 323 |
| Gauge | 15 | 32 | 41 | 180 | 300 |
| Occupancy (median number of occupants / Gauge) | 21 % | 32 % | 40 % | 63% | 102 % |
| Session duration (min) | 75 | 90 | 120 | 120 | 240 |
| **Parameter of CO<sub>2</sub> distribution in the observed sessions** |  |  |  |  |  |
| 3rd quartile CO<sub>2</sub> (ppm) | 571 | 955 | 1188 | 1434 | 2152 |
| max CO<sub>2</sub> (ppm) | 658 | 1056 | 1292 | 1697 | 2344 |
| **Mechanical ventilation performance in the observed sessions** |  |  |  |  |  |
| clean air flow per occupant at 100% occupancy (m<sup>3</sup>/ h / occupant) | 3 | 9 | 15 | 24 | 91 |
| Percentage of clean air flow per occupant at 100% occupancy, with respect to the legal airflow of 25 m<sup>3</sup> / h / occupant | 13 % | 35 % | 60 % | 94 % | 363% |

In 47 out of the 57 classroom sessions (82%), the clean air flow at 100% occupancy was estimated below the legal threshold of 25 m^3^/h/occupant at 100% occupancy. In 32 out of the 41 rooms (78%), at least one session led to an estimation of a clean air flow at 100% occupancy below 25 m^3^/h/occupant. Figure 3 shows a typical measurement in a 300-seat, 1,360 m^3^ amphitheatre, occupied to 32% of gauge.

**Figure 3.**
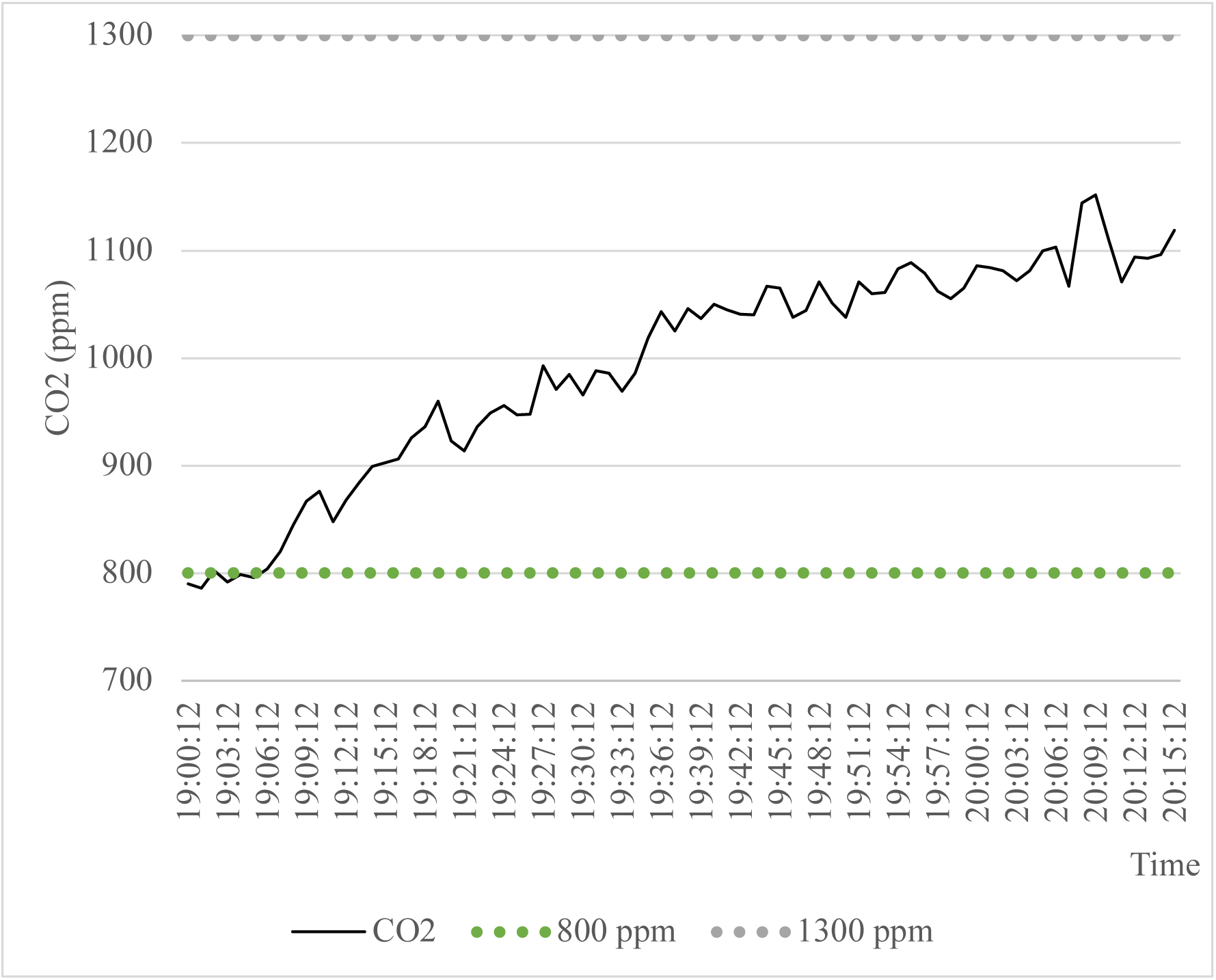
Typical CO_2_ profile in room Ta, a room of 1,360 m^3^ occupied by 95 people for a gauge of 300 (32% occupancy). 3^rd^ quartile of CO2 = 1071 ppm. Clean air flow during session = 35 m^3^/h/occupant. Clean air flow at 100% occupancy = 11 m^3^/h/occupant. Percentage of gauge reduction required to meet legal airflow of 25 m^3^ / hour / occupant at 100% occupancy = 55% (78% to reach the 50 m^3^/h/occupant target of HCSP).

#### 3.4.2 Study of CO_2_ decay data after room use

The decay of the CO₂ concentration was studied subsequent to the cessation of room use and the closure of all apertures in nine rooms (including one, the Ne amphitheatre, before and after repair of the air handling unit). The results obtained are presented in Table 4. Figures 4 and 5 show the decay curves for amphitheatre Ne, before and after repair of the air handling unit.

**Figure 4.**
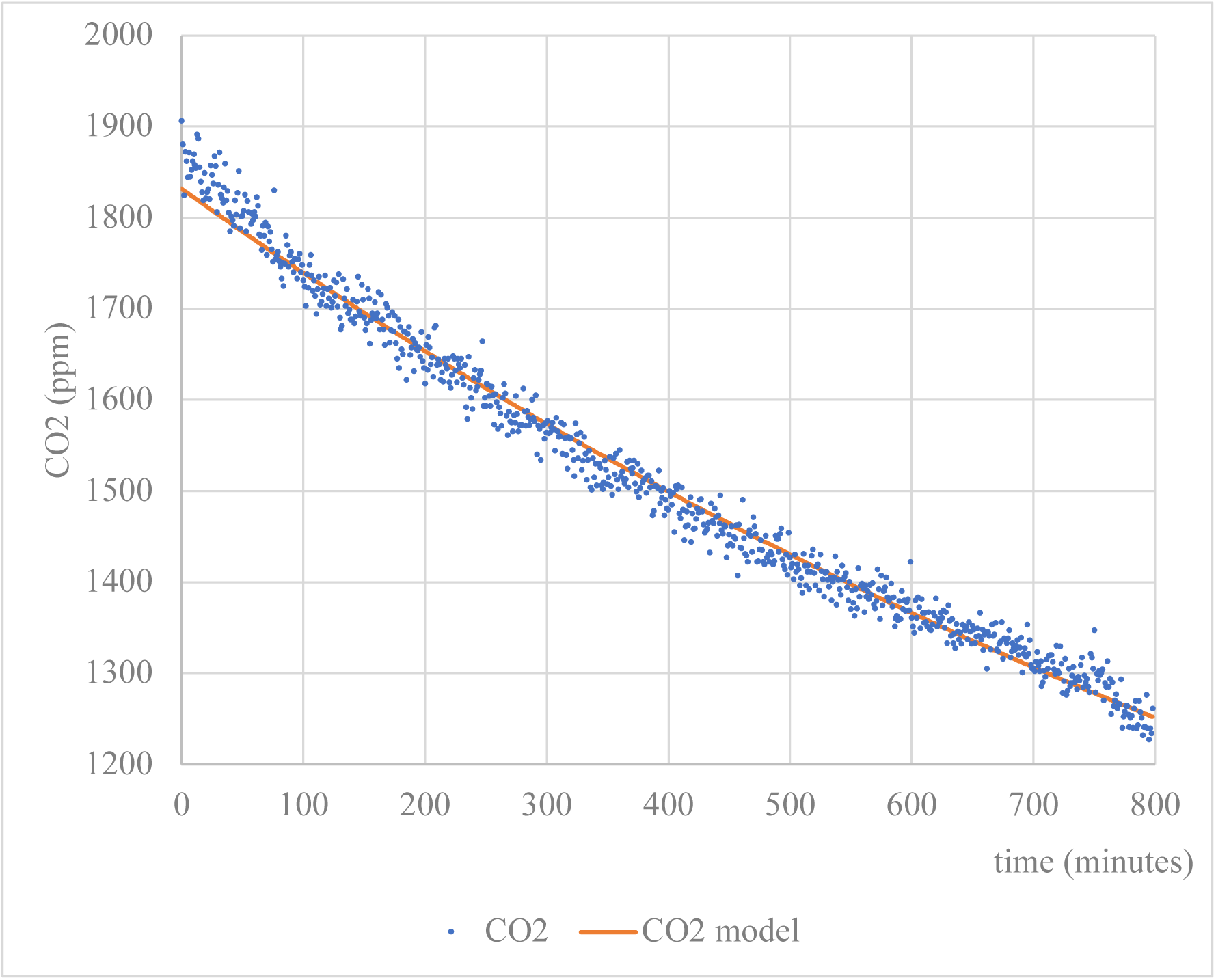
Model of CO_2_ removal in room Ne, a room of 1,040 m^3^ with a gauge of 240 (room emptied after use, all doors and windows closed) during one night (800 minutes). r = 0.994. Renewal rate = 0.04 volume/h. Clean air flow per occupant at 100% occupancy = 0.2 m^3^/h/occupant (< 1% of the regulatory floor).

**Figure 5.**
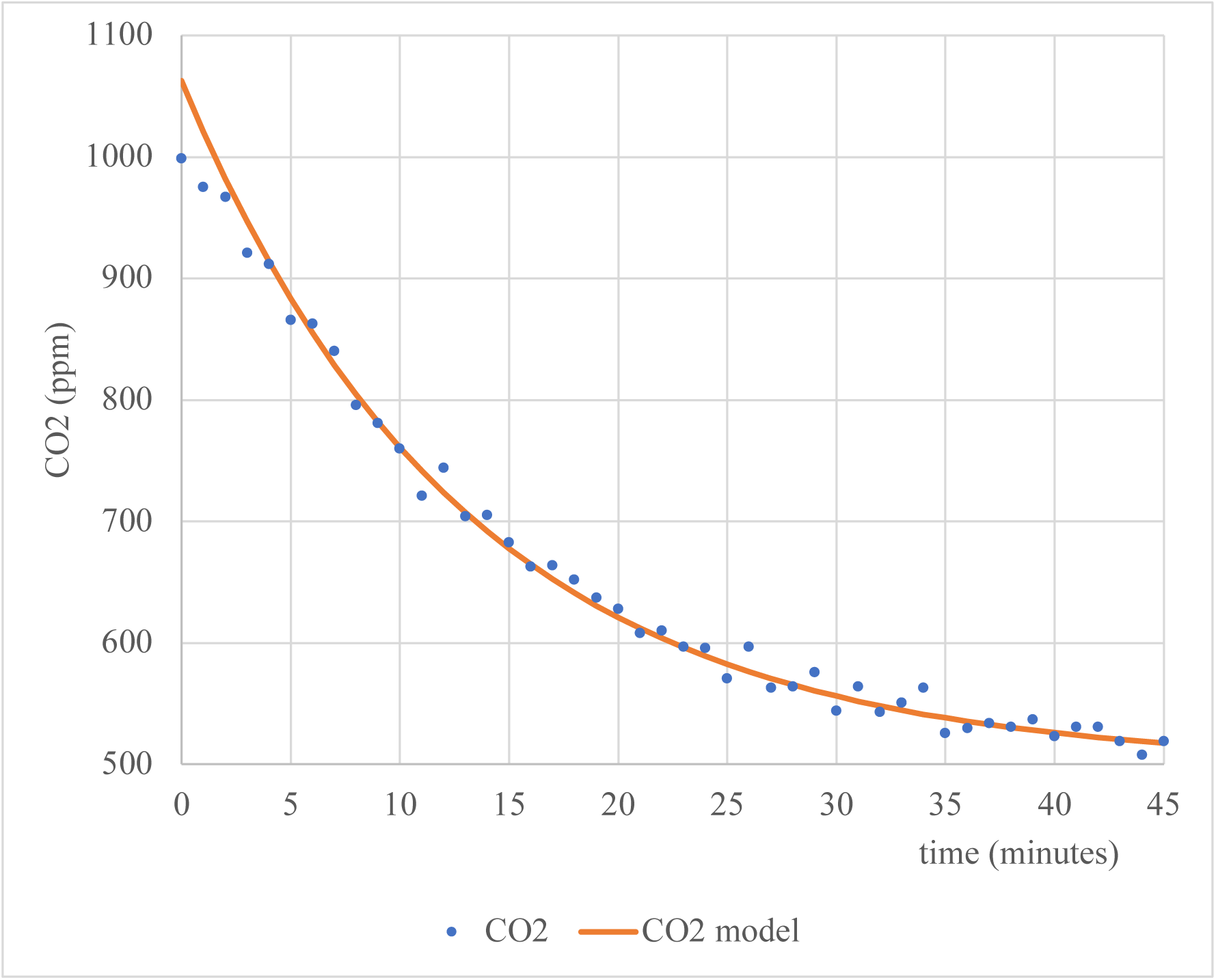
Model of CO_2_ removal in room Ne, a room of 1,040 m^3^with a gauge of 240 (room emptied after use, all doors and windows closed) during 45 minutes. r = 0.996. Renewal rate = 4.6 volume/h. Clean air flow per occupant at 100% occupancy = 20 m^3^/h/occupant (80% of the lower legal limit). *Note: This measurement was made after the air handling unit was repaired*.

**Table 4.** Summary of ventilation performance estimates (sorted by portion of legal airflow per occupant at 100% occupancy) based on analysis of CO_2_ decay after evacuation of the amphitheatres and closure of openings.

| Build ing | Ro om | Ga uge | Volu me (m<sup>3</sup>) | Correl ation coeffici ent | Renewal rate (volume / hour) | clean air flow (m<sup>3</sup> / hour / occupant) at 100% occupancy | portion of legal airflow per occupant at 100% occupancy |
| --- | --- | --- | --- | --- | --- | --- | --- |
| N | e | 240 | 1040 | 0.994 | 0.04 | 0.2 | 1% |
| T | b | 125 | 377 | 0.976 | 0.3 | 0.9 | 4% |
| N | b | 240 | 1040 | 0.990 | 0.84 | 3.6 | 14% |
| L | f | 176 | 900 | 0.995 | 0.73 | 3.7 | 15% |
| G | f | 122 | 390 | 0.982 | 1.4 | 4.5 | 18% |
| T | a | 300 | 1360 | 0.990 | 1.1 | 5.0 | 20% |
| T | c | 125 | 434 | 0.990 | 2.2 | 7.6 | 30% |
| P | a | 72 | 470 | 0.985 | 1.8 | 11.8 | 47% |
| N | e | 240 | 1040 | 0.996 | 4.6 | 19.9 | 80% |
| W | a | 120 | 710 | 0.991 | 3.5 | 20.7 | 83% |

### 3.5 Compliance with legal requirements and HCSP recommendations in the rooms surveyed by the UGA sensors

Over the period from September 2022 to June 2023, the UGA deployed 117 CO_2_ sensors in 101 rooms. Of these 101 rooms, 23 had concentrations greater than 5,000 ppm (15 in rooms without mechanical ventilation, 8 in rooms with mechanical ventilation). Figure 6 shows the peaks of CO_2_ above 2,500 ppm observed in one of these rooms.

**Figure 6.**
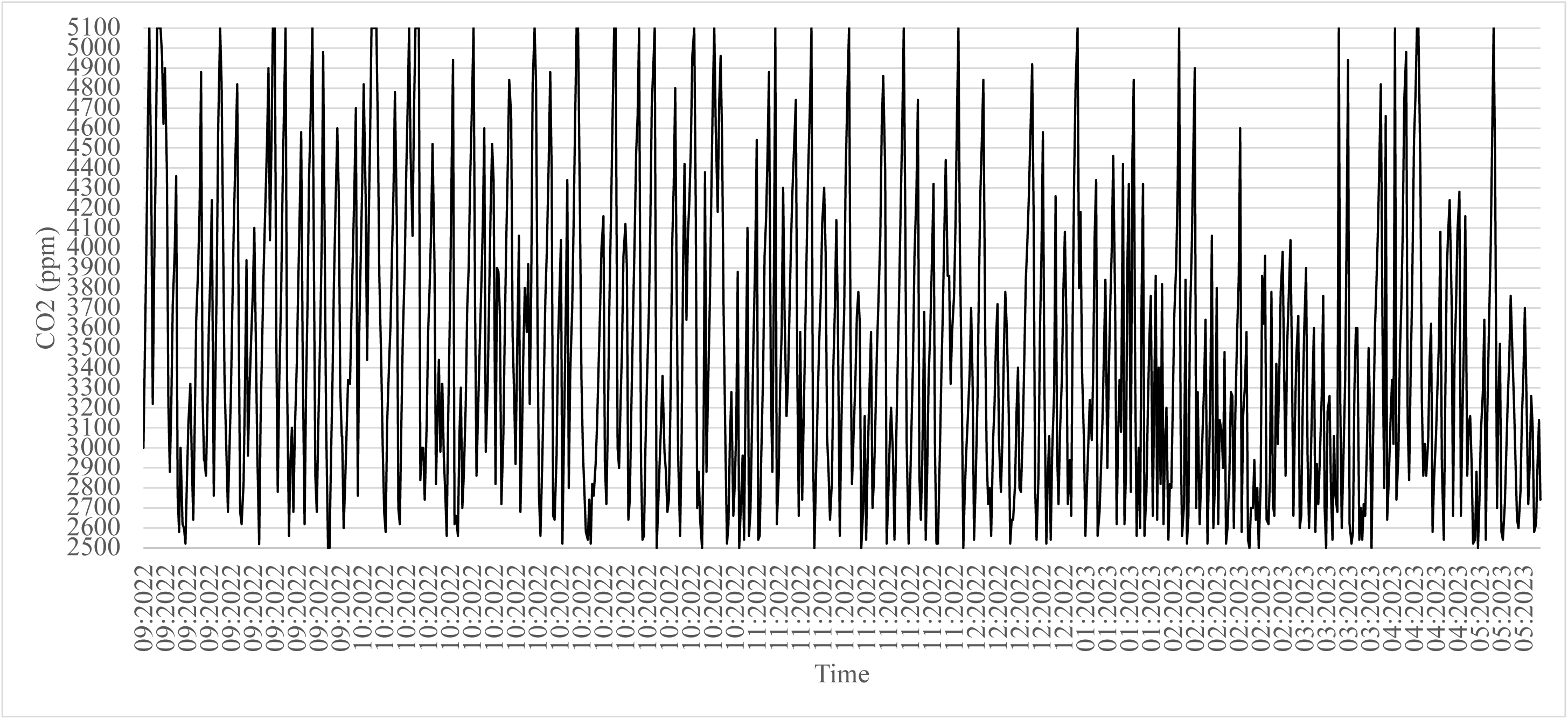
94 CO2 peaks above 2,500 ppm (28 of which above 5,000 ppm) were observed between September 2022 and June 2023, in one of the 23 UGA classroom (out of 101 monitored) where peaks above 5,000 ppm were measured (only values above 2,500 ppm were plotted in this graph). This room has a gauge of 44 occupants, an area of 51.8 m2, and a volume of 140 m3 (3.2 m3/occupant).

Over the period from 26 March to 16 April 2024 (3 weeks), 5 lecture theatres (i.e. 31% of the 16 lecture theatres with more than 100 seats where the UGA had installed sensors) had CO_2_ peaks above 2,500 ppm. Room Xa was a room where very high CO_2_ concentrations were observed most frequently and was therefore monitored more carefully. Figure 7 summarizes the CO_2_ peaks observed during the occupation of room Xa in a 4 days week, from April 29 to May 2 (May 1^st^ is a holiday in France).

**Figure 7.**
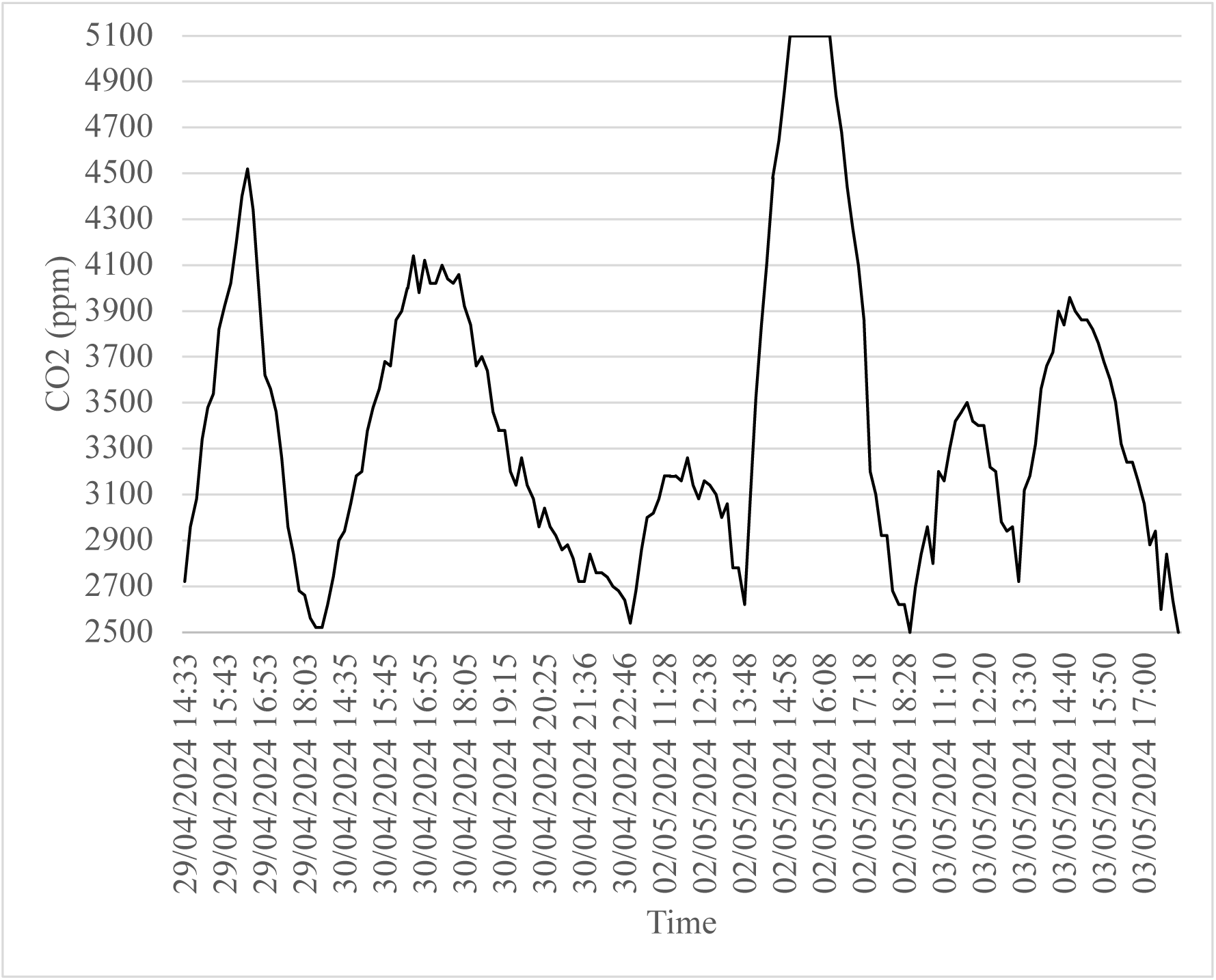
Summary of the 6 CO_2_ peaks above 2,500 ppm observed in room Xa in a 4 working days week (from March 29 to May 2, 2024). Only values above 2,500 ppm were plotted in this graph. This room has a gauge of 154 occupants, an area of 128 m^2^, and a volume of 434 m^3^ (2.8 m^3^/occupant). See photos of this room in Figure SM.1.

By studying the decrease in CO_2_ levels after the CO_2_ peak, it was possible to estimate their clean air renewal rates, and where the volume could be estimated, their clean air flow per occupant at 100% occupancy (Table 5).

**Table 5.** Clean air renewal rate and clean air flow per occupant at 100% occupancy in 5 lecture theatres with more than 100 seats where the UGA had installed sensors and where peaks above 2,500 ppm were observed. Clean air flow was estimated by modelling the decay of CO_2_ concentrations.

| Building | Room | Gauge | Volume (m<sup>3</sup>) | Renewal rate (volume / hour) | clean air flow (m<sup>3</sup> / hour / occupant) at 100% occupancy | portion of legal airflow per occupant at 100% occupancy |
| --- | --- | --- | --- | --- | --- | --- |
| V | C | 335 | 900 | 0,12 | 0,32 | 1% |
| O | B | 108 | 337 | 0,15 | 0,47 | 2% |
| V | B | 348 | 900 | 0,2 | 0,52 | 2% |
| X | a | 154 | 434 | 0,5 | 1,41 | 6% |
| W | A | 124 | 631 | 1,4 | 7,12 | 28% |

In one particular case, a long CO_2_ plateau at over 5,000 ppm was observed. In this case, it was possible to use the CO_2_ decay model to estimate that the maximum concentration of CO_2_ to which the occupants of this lecture theatre were exposed was between 8,840 ppm and 10,608 ppm, depending on the actual end time of the examination, which was scheduled for 4:30 pm, but may have been delayed (Figure 8).

**Figure 8.**
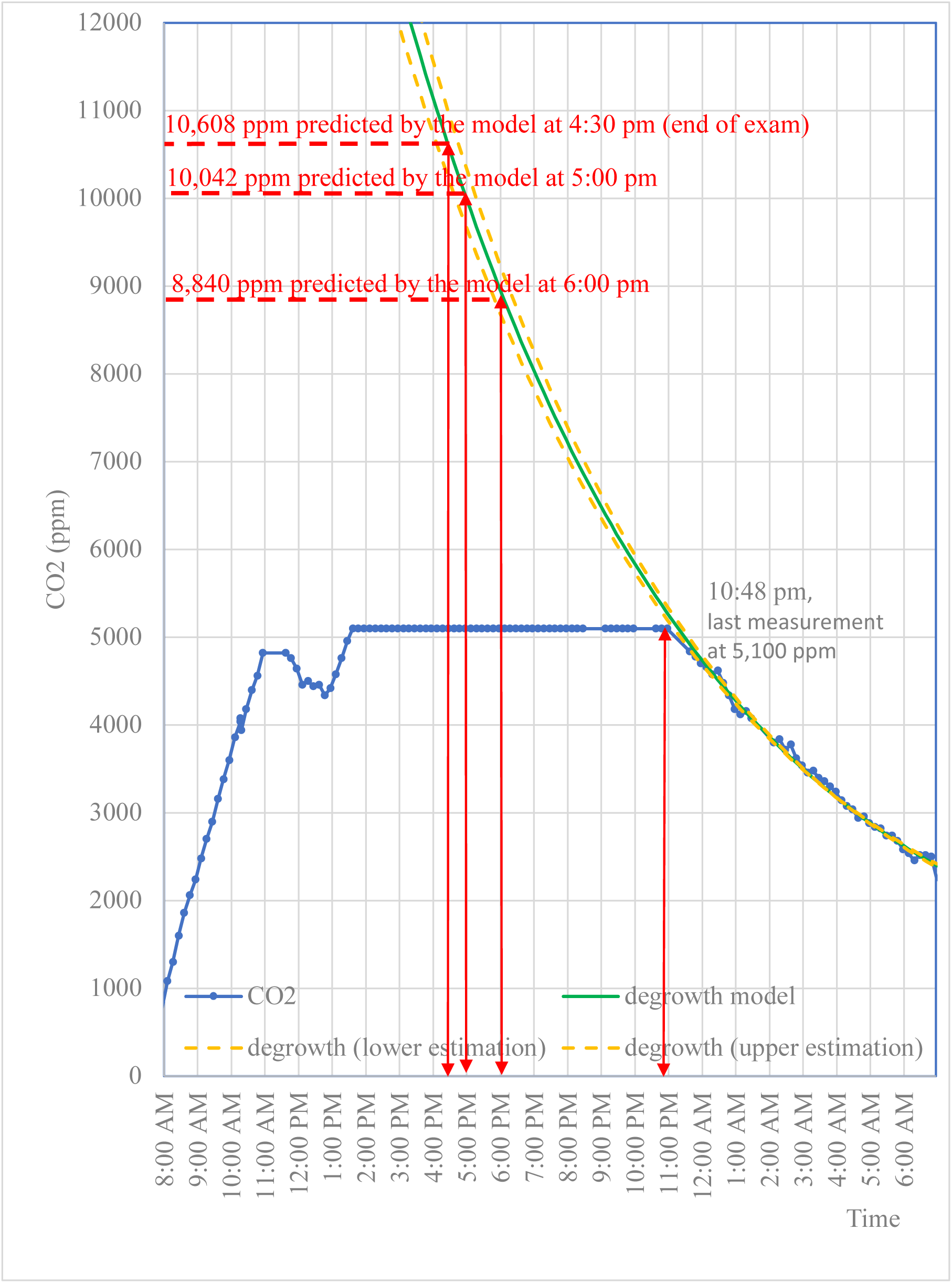
Growth, then degrowth of CO_2_ levels in room c of building V (area = 255 m^2^, volume = 990 m^3^, gauge = 335). The upper limit of the sensor is 5,000 ppm. Degrowth model is estimated from 22:58 to 6:59 (43 measurements, r = 0.997). The dotted lines correspond to the lower and upper limits of the 95% confidence interval.

## 4 Discussion

### 4.1 Applicability of the proposed method to identify dangerous situations and to propose practical recommendations

#### 4.1.1 Involvement of the lecturers

The involvement of the lecturers made it possible to collect precise data on the number of occupants and the condition of the openings (all the measurement files sent were accompanied by the required details on this point). The lecturers taking part in the study reported no difficulties in collecting this information, which is a major advantage over processing data from fixed sensors installed in the rooms, with no information on the number of occupants or the state of the openings. Using the mobile CO_2_ sensors made available to the teachers proved very easy, with all of them having no difficulty installing the necessary application on their mobile phones. Transferring the measurements and information about the measurement conditions to the cloud made available to them posed no particular problem either. The study also raised participants’ awareness of the need for better ventilation in premises with insufficient natural ventilation, and of the reality of the problem of insufficient mechanical ventilation in many rooms.

#### 4.1.2 Uncertainty of the estimation of the indicators of air quality from our measurements

As described in sections 2.5.1 and 2.5.2, the clean air flow is estimated on the basis of CO_2_ measurements, using the classical model of CO_2_ concentration in a well-mixed space [19]. CO_2_ measurements are known with an uncertainty of ± (30 ppm + 3% of reading), which corresponds to ± 5% ppm for a concentration of 1,500 ppm (and between 3 and 5% for higher concentrations).

For estimations of air flow based on CO_2_ measurements while the room is occupied, the CO₂ growth model operates under the assumption of homogeneous mixing of the air within the room. However, it is well known that CO_2_ concentrations can vary from one point of the room to another, depending on natural convective air movements [22], especially when an air current is established after opening windows or doors, or on the proximity of room occupants [23]. These differences can reach 10% of measurements [24]. Therefore, the optimal scenario would involve the strategic placement of multiple CO2 sensors throughout the room, whereas only a single sensor was utilized per session of measurements. Given the utilization of a single sensor in each session, the total uncertainty in the CO2 measurements was considered to be ±15% (5% due to the sensor uncertainty, 10% due to the inhomogeneity).

In order to estimate the limit concentration of CO_2_, that was reached at steady state, the third quartile *Q_3_* of CO_2_ measurements obtained during a session of room use was utilized in Equation (4) – rather than the maximum – for two reasons. Firstly, this approach excluded potential outliers. Secondly, this deliberate overestimation of the clean air flow ensured that rooms considered incompatible with the law truly were. This approach was also adopted to determine the outdoor concentration of CO_2_, which was set at 500 ppm, representing the highest concentration measured. The utilization of *Q_3_* as opposed to the maximum of CO_2_ offers a distinct advantage, namely the fact that the associated uncertainty is considerably less pronounced in comparison to the uncertainty inherent in discrete CO_2_ measurements. Nonetheless, the estimation of the uncertainty on *Q_3_* will be maintained at 15%. Given that Equation (4) utilized to estimate the clean air flow we use model is *D_N_ = 20,200/(Q_3_-500)*, an uncertainty of ± 15% in *Q_3_* yields an uncertainty of *D_N_* inferior to 22.5% when *Q_3_* ≥ 1,500 ppm.

Another source of uncertainty comes from estimating the average CO_2_ generation rate by the human beings present in the room. The value of 5.6 mL/s (or 20.2 L/h) was utilized, as proposed by the Air Infiltration and Ventilation Centre [20]. This value is compatible with the inhalation rates used by [25] for different activity levels (0.49 m^3^/h for resting, 0.54 m^3^/h for standing), which, taking into account an average CO_2_ concentration of 4% [26] correspond to 19.6 L/h and 21.6 L/h, respectively. However, other publications suggest lower CO_2_ generation rates. For example, [27] suggests an average value of 16.2 L/h for a classroom of 64 students and one teacher, and [28] considers that an activity of “sitting reading, writing, typing” corresponds to a metabolism of 1.3 times the basal metabolism, corresponding to an average value of 18.0 L/h. Once more, we elected to overestimate the clean air flow, which consequently guided our decision to utilize the estimate proposed by [20].

The variability of estimates of average human CO_2_ production rates underscores the importance of preferring the CO_2_ decay method. In addition, during decay measurements, the risk of mixture inhomogeneity is reduced because all openings are closed and the room is empty, so the uncertainty is also reduced with this method. Finally, the fact that the model is fitted to all measurements also reduces the uncertainty. This explains why the confidence interval of the decay model is small, as can be seen in Figure 8. For all these reasons, this method should be used whenever possible.

#### 4.1.3 Reliability of the estimation of the indicators used to characterize violations of legal requirements for air quality

Analysis of the estimated clean air flows shows that an underestimation of 22.5% of the clean air flow would lead to classifying 24 instead of 32 (59% instead of 78%) of the 41 rooms with mechanical ventilation as uncompliant with the legal minimum of 25 m^3^/h/occupant. Such an error would have no practical consequences. In fact, universities do not have the resources to immediately address all air handling unit deficiencies in more than half of their mechanically ventilated rooms, so an error in the classification of 20% of the rooms will not have major consequences: university administrators will have to work first on improving their worst rooms in terms of air quality.

#### 4.1.4 Comparison of flow estimates during room occupancy and CO_2_ decay

In rooms with mechanical ventilation, where the clean air flow per occupant could be estimated both by estimating the limit concentration of CO_2_ at steady state and by modelling the decay of CO_2_ after evacuation of the room and closure of the openings, the estimate obtained by the first method was systematically higher than that obtained by the second. The first reason for this difference is that steady state is often not reached before the end of the session. A second reason is the deliberate choice of the 3^rd^ quartile of CO_2_ concentrations as the limit value, which was justified in the previous section. Finally, in certain circumstances, opening of doors and windows provides more clean air than mechanical ventilation. It is therefore not surprising that the estimate obtained after evacuating the room and closing the openings is systematically lower than that obtained in real conditions, where the openings can be opened. This is illustrated by the example of room Ne, where the clean air flow was estimated during 6 classroom sessions. The 6 estimates of clean air flow (in m^3^/h/occupant at 100% occupancy) for this room are 3.5, 4.0, 4.3, 4.6, 5.4, 8.1. The relative homogeneity of the first 5 estimates should be noted, with the estimate of 8.1 m^3^/h/occupant appearing to be an outlier. These measurements were obtained when the mechanical ventilation in this 240-seat auditorium was not operational. This auditorium has the unusual feature of having 3 doors, which were open (two main doors at the bottom of the amphitheatre, opening onto a patio where a large number of students may walk, and where the CO_2_ concentration can therefore vary significantly, and a third at the very top, opening onto a corridor). A slight draught was felt by the lecturer at the bottom of the amphitheatre. The estimated clean air flow per occupant for this same room using the decay method (Figure 4) is 0.2 m^3^/h/per occupant at 100% occupancy. It can be seen that opening the doors provides a significant flow of clean air, although it is far from sufficient.

Estimating the flow of clean air at 100% occupancy using the CO_2_ decay method after evacuating the room and closing the doors proved particularly simple to implement. The first tests of this method were carried out on the Ne room, when its mechanical ventilation was down. As a result, the sensor had to be left in place overnight to observe a sufficient drop in CO_2_, which posed practical constraints when it came to retrieving it the following morning. But when the mechanical ventilation is working more or less correctly, measurements over an hour or so after the doors have been closed are sufficient. However, it is difficult to ask a lecturer to stay for an hour after the end of the lesson. Extending this method to all classrooms would therefore require institutional mobilisation, with the involvement of the building maintenance staff.

For rooms with mechanical ventilation, this method of modelling CO_2_ decay seems the simplest to implement, for rapidly characterising all the rooms with mechanical ventilation in a university. By simply taking this measurement after the end of the last lesson of each day, it is possible with one sensor to characterise 20 rooms in one month, i.e. around 20 working days. For an investment of around €2,000, or 10 sensors, 200 rooms could be characterised each month, at the cost of a moderate human investment (a maximum of ten minutes to transmit and process each series of measurements characterising a room). However, this requires institutional mobilisation and political will.

### 4.2 Detecting situations likely to endanger the health of users and alerting the university administration

Well before the launch of the participatory campaign, several warnings about the poor quality of the air in the university had been issued, including warnings about risks to the health of staff and students, without any solutions to the problems identified being implemented. The participatory campaign was launched to assess the scale of the problem across the whole university.

#### 4.2.1 Risk situations in rooms without mechanical ventilation

Table 1 shows that none of the studied rooms meets the legal limit of at least 15 m^3^/occupant. Of the 14 rooms without mechanical ventilation in our study, where the volume was known, to meet this minimum:

- 2 rooms (14%) should reduce their gauge by less than 50%,

- 6 rooms (43%) should reduce their gauge by between 50 and 70%,

- 6 rooms (43%) should reduce their gauge by more than 70%.

As early as December 2023, the participatory campaign demonstrated that failure to comply with the minimum volume per occupant in rooms without mechanical ventilation led very quickly (typically within 15 minutes), when the openings (doors and windows) were closed, to the threshold of 1,300 ppm being exceeded, with regular measurements of over 4,000 ppm, and even 5,000 ppm, as Table SM.1, Table 2, and Figures 1, 2 and 6 amply demonstrate. Figure 7 illustrates the frequency with which 2,500 ppm concentrations are exceeded in a 154-seat amphitheatre (2.8 m^3^/occupant) without mechanical ventilation.

Of course, the method used in no way allows us to claim that the 15 rooms without mechanical ventilation studied by the participatory measurement campaign are representative of the situation in all the university’s rooms. They were not chosen at random; these were the rooms where the campaign volunteers taught. However, the observations of the participatory campaign are compatible with the analysis of the data from the fixed CO_2_ sensors. It is very worrying to note that, as table 2 shows, in 52% of cases, the first quartile of CO_2_ concentrations measured in these rooms exceeded the threshold of 1,300 ppm (and 97% of this first quartile exceeded the threshold of 800 ppm recommended by the HCSP). These rooms, which have no mechanical ventilation, are usually of relatively limited gauge (typically designed for 30 to 40 occupants, and are usually classrooms for tutorials or practical work). However, there is at least one case where a 154-seat lecture theatre has no mechanical ventilation. This is room Xa, where the volume per occupant is 2.8 m^3^/occupant, or 19% of the legal minimum threshold of 15 m^3^/occupant (see Figure SM.1 for photos and plans of this room). Under these conditions, it is not surprising that plateaus of over 5,000 ppm for more than an hour are regularly recorded there, even with an occupancy rate of less than 50%. Such levels have been observed in particular during examination periods. This is particularly problematic in view of publications showing a decline in psychomotor and cognitive abilities at concentrations above 1,000 ppm, and especially above 2,500 ppm [29] [10], as well as an increase in the rate of headaches [10] [12].

Our campaign therefore suggests that any room without mechanical ventilation that does not meet the threshold of 15 m^3^/occupant should be considered hazardous to the health of occupants, as long as the openings cannot be permanently opened when the room is in use.

#### 4.2.2 Risk situations in rooms with mechanical ventilation

Here again, our methodology cannot guarantee that all 41 rooms in 15 of the university’s buildings are representative. However, it is noteworthy that if the legal minimum of clean air flow of 25 m^3^/h/occupant is considered the objective, a mere nine of the 41 rooms examined (22%) would not necessitate a reduction in gauge. Conversely, if the objective is to adhere to the most recent HCSP recommendation (a minimum clean air flow of 50 m^3^/h/occupant, corresponding to a maximum CO_2_ concentration of approximately 800 ppm), it is observed that merely two of the 41 rooms meet this criterion (with doors of these two rooms open to the outside during the session). Consequently, in order to adhere to the recommendations outlined by the HCSP, it is imperative that at least 39 of the 41 rooms under consideration undergo a reduction in gauge, with 35 of these rooms required to undergo a reduction greater than 70%. It should also be remembered that the estimates of clean air flow during classroom sessions are deliberately “optimistic” for the reasons discussed in section 4.1.2, and that measuring the flow of clean air per occupant using the CO_2_ decay method would certainly give even more drastic estimates of the reduction in gauge.

#### 4.2.3 The university’s ability to respond to air quality alerts

The university has two levers at its disposal to reduce the risk to which its staff and students are exposed as a result of poor indoor air quality. The first of these levers is technical, and can be activated when the problem arises from a malfunction in the air handling units. This was the case for the Ne amphitheatre. When the relevant university departments intervened, it was found that the air handling unit in this amphitheatre had stopped working. Simply restarting it brought about a considerable improvement in the flow of clean air per occupant, which rose overnight from 0.2 m^3^/h/occupant to 20 m^3^/h/occupant (80% of the legal minimum of 25 m^3^/h/occupant, and 40% of the HCSP recommendation of 50 m^3^/h/occupant).

The second lever is political and organisational. The university needs to become aware of the seriousness of the public health problem posed by the quality of its indoor air. In reality, despite explicit advice from the country’s highest public health authority, the HCSP, the message is struggling to get through. Such awareness would provide the means to detect malfunctions in the air handling units and correct them where possible, as well as defining a policy for adapting the use of rooms, which could be used to improve indoor air quality.

The fixed sensors deployed by the university have the advantage of constantly monitoring the CO_2_ concentration in the rooms equipped with them. This is very useful for detecting malfunctions in the air handling units. However, during the course of our study, no one was responsible for sounding the alarm if the thresholds were exceeded. Thus, it was within the framework of the study described here that analysis of the CO_2_ signal recorded continuously in the Vc amphitheater (see Figure SM.2 for photos and plans of this room) made it possible to detect the shutdown of the air handling unit in this building, the consequences of which are summarized in Figure 8. This shutdown during an examination led to considerable exceedances of the legal thresholds, with probably a peak of CO_2_ concentration around 10,000 ppm. Such levels are utterly unacceptable, particularly during an examination period when students are mobilizing all their cognitive capacities. It has previously been noted that these capacities are diminished above 1,000 ppm, and more specifically above 2,500 ppm. At 10,000 ppm, the risk of contamination by airborne infectious agents is considerably increased (21% of the air breathed in comes from the occupants’ lungs). But direct toxicity must also be considered. This is characterized by the Occupational Exposure Limit (OEL), which must not be exceeded for 8 hours (OEL 8h) or for 15 minutes (Short Term OEL). The 8h OEL for CO_2_ is 5,000 ppm [30]. Short Term OEL values vary from country to country (10,000 ppm in Germany, 30,000 ppm in the USA, France has not defined one). The concentrations shown in Figure 8 are therefore particularly problematic.

It should also be possible to put in place a suitable organisation to prevent the recurring risks of CO_2_ limits being exceeded. Once again, the example of the Vc amphitheatre is typical. In building V, the air handling unit stopped working due to a malfunction in the dialogue between this unit and the fire detection system. This dialogue regularly leads to the air handling unit shutting down, which can only be restarted by the intervention of a member of staff. Despite repeated warnings, it has not been possible to arrange for the CO_2_ concentration in the two lecture theatres in Building V with fixed CO_2_ sensors to be read at least daily. This is how the incident summarised in Figure 8 occurred, even though reading the signals from these sensors several days before the examination summarised in Figure 8 would have made it possible to detect the shutdown of the air handling unit and therefore to restart it before the examination that led to the 10,000 ppm peak. Several weeks before the incident summarised in Figure 8, the authorities had been asked to put in place a procedure to ensure that the CO_2_ sensors in this building were checked twice a day and that action was taken immediately to restart the air handling unit in the event of an unexpected shutdown. The alert had even been given 48 hours before the examination summarised in Figure 8. No measures were taken to protect the occupants. Worse still, although a specific alert had been filed following this incident on the official register, a new similar incident occurred a few weeks later, with the 5,000 ppm limit value again being exceeded during an examination.

The university also needs to organize itself to react to a known systemic risk. Signals showing the reality and seriousness of the problem in a large proportion of the rooms at UGA had been available at the highest levels of the university since July 2022. The poor quality of the indoor air was discussed on numerous occasions by the university’s specialist risk management bodies, without appropriate corrective or preventive action being systematically taken in line with the HCSP recommendations for CO_2_ levels in excess of 1,500 ppm. Figures similar to those in Figure 6 have been discussed several times at the highest level, without the university’s website mentioning the existence of the problem, and without appropriate warning measures being systematically taken (for example, during months, most of the rooms concerned did not display a poster warning of the existence of a danger, encouraging users to open the openings as much as possible, asking people entering the room to use a surgical mask at the slightest sign of respiratory infection, and suggesting that immuno-compromised people protect themselves). No decision was taken to reduce the gauge of the rooms, even in the face of major inadequacies. Amphitheatre Xa, for example, continued to be used to accommodate students in conditions that regularly led to considerable breaches of the legal thresholds (up to more than 5,000 ppm, see Figure 7), even though the failure to comply with the minimum threshold of 15 m^3^/occupant for this room without mechanical ventilation should have led to the 154-seat gauge being reduced to 29.

Finally, when a malfunction is reported in a particular room, it is important to organise a rapid response. The university has a specific register in which its staff can officially report any risk to their health or that of their colleagues and students. In our experience, this register did not allow for rapid reactions when exceedances of the legal CO_2_ thresholds were reported. For example, an initial informal alert following a CO_2_ measurement in lecture theatre Ne was issued in January 2022. As this alert did not lead to any action being taken, a second alert was filed on the ad hoc register in January 2023, which was also not acted upon. A third alert was filed in January 2024. This alert was initially closed by the administration, which considered that the problem was already known. The staff member concerned had to challenge this decision to close the alert before the university’s health and safety committee. A specialist was finally sent to check the operation of the air handling unit, found that it had been shut down, and restarted it. This highlights how difficult it is for a university to listen to the alarm signals it receives.

### 4.3 Practical recommendations to the administrators of universities regarding indoor air quality

The findings of this study, delineated in the preceding subsections, can be extrapolated to formulate recommendations for university administrators concerning indoor air quality. In certain circumstances, the implementation of a constant air current throughout the duration of room utilization can become impractical. These circumstances include instances where the outside temperature is excessively low or high, or where there is an excessive amount of noise. In such situations, all rooms lacking mechanical ventilation systems should be regarded as potentially hazardous, as their air quality is likely to be substandard unless adequate ventilation is ensured by opening doors and windows between classes. To address this concern, it is recommended that each room be equipped with a sign instructing students and faculty to open windows and doors for at least five minutes between classes. The volumes and dimensions of all affected rooms should be documented for future reference. The university should define a strategy to reorganize courses so that eventually no course will be held in a room with less than 15 m^3^/occupant.

The clean air flow of all rooms with mechanical ventilation should be estimated at least once a year, preferably using the CO_2_ decay model. This can be organized at relatively low cost by applying the principles of the “participatory campaign” used in this study. All air handling units that cannot guarantee at least 25 m^3^/h/occupant should be checked immediately and repaired if possible. In a first phase, the university should define a strategy to reorganize courses in such a way that eventually no course will be held in a room with a clean air flow of less than 25 m^3^/h/occupant (in a second phase, the target should be set at 50 m^3^/h/occupant).

Defining these strategies to improve the indoor air quality in universities is all the more important in countries where legislation is relatively less protective. French legislation is currently one of the least protective in Europe in terms of minimum ventilation rates (25 m^3^/h/occupant in France, while the minimum legal value in Europe is 20 m^3^/h/occupant and the maximal legal value is 90 m^3^/h/occupant) [31]. As was already mentioned, HCSP recommends to target a maximum concentration of CO_2_ of 800 ppm (while the legal threshold is 1,300 ppm), and a minimum flow of clean air of 50 m^3^/h/occupant (while the legal threshold is 25 m^3^/h/occupant). Universities should therefore prepare to upcoming more demanding legal prescription in terms of Interior Air Quality.

## 5 Conclusion

This study has shown that it is useful and easy to mobilize teachers to measure CO_2_ concentrations in the rooms of a university, in order to characterize risk situations. The study shows that rooms without mechanical ventilation are hazardous to the health of their occupants when they are used with openings closed and without complying with the legal minimum volume of 15 m^3^/occupant. For rooms without mechanical ventilation, the study confirms that clean air flows below the legal threshold of 25 m^3^/h/occupant are insufficient to guarantee the quality of indoor air, and therefore the health of their occupants. The finding that the majority of the rooms under study did not comply with the legal requirements suggests that this problematic situation may also be present in a significant proportion of rooms at the university surveyed. Our study demonstrated the ease and simplicity of implementing the method for characterizing the ventilation performance of an amphitheater, which consists of measuring the decrease in CO_2_ after the amphitheater has been evacuated and the doors closed. This method could be implemented very quickly and easily to characterize the ventilation performance of all the rooms in the university that have mechanical ventilation. With the political will of the administration, it would be possible to characterize the ventilation in all these rooms in just a few months. However, compliance with the legal threshold of 25 m^3^/occupant/h, and even more so with the target of 50 m^3^/occupant/h, will necessarily involve substantial investment and will take time. In the meantime, it will be necessary to work on the use of the most problematic rooms, so as to be able to reduce their gauge. Given the university’s shortage of rooms, this is a considerable challenge. To address this issue, it is essential to quantify the problem precisely. The method proposed here is a promising approach for achieving this objective.

## Supporting information

Table SM.1. Summary of measurements in the 15 rooms without mechanical ventilation

Figure SM.1 Photos and dimensions of room Xa, a room without mechanical ventilation where concentrations above 5,000 ppm were frequently recorded

Figure SM.2 Photos and dimensions of room Vc, where the air handling unit malfunctioned, resulting in CO2 concentrations above 10,000 ppm

## Data Availability

The data used to support the findings of this study are available from the corresponding author upon request.

## Acknowledgments

This study would not have been possible without the involvement of the SNESUP-FSU union, who funded the sensors and advertised the study, and without the involvement of 20 lecturers and researchers from Grenoble Alpes University, who volunteered for the participatory campaign described here: we would like to extend our warmest thanks to them. The author is grateful to Romain Guichard (Laboratoire d’Aéraulique, VentilAtion, Thermique et qualité d’AiR, INRS) for his advice on modelling CO_2_ concentrations in rooms and for his careful proofreading of the article, as well as to the members of the CO_2_ project https://projetco2.fr/ (Pascal Morenton, Benoît Semin) for advice on CO_2_ measurement methods and for certain bibliographical references.

## Concise description of Supplementary figures and tables

**Table SM.1.** Summary of measurements in the 15 rooms without mechanical ventilation.

**Figure SM.1** Photos from the front and rear, and dimensions, of room Xa. This is a room without mechanical ventilation where CO_2_ concentrations above 5,000 ppm were frequently recorded. This room has a gauge of 154 occupants, an area of 128 m^2^, and a volume of 434 m^3^ (2.8 m^3^/occupant). The lower windows cannot be opened. The upper windows can only be partially opened, so that the maximum opening area of the windows is 2.7 m^2^.

**Figure SM.2** Photos from the front and rear, and dimensions, of room Vc. This room with mechanical ventilation had problems with its air handling unit, leading to concentrations of CO_2_ above 10,000 ppm. This room has a gauge of 335 occupants, an area of 255 m^2^, and a volume of 990 m^3^.

