## Supplementary material for "Indoor air quality at a French university: *a participatory CO2 measurement campaign highlights the wide gap between reality and the law*": Table SM.1. Summary of measurements in the 15 rooms without mechanical ventilation

| **Building** | **Room** | **Session Id** | **Gauge** | **Median number of occupants** | **Session duration (min)** | **3rd quartile CO2 (ppm)** | **Max CO2 value (ppm)** | ***$J_{HCSP}$*** Recommended gauge to meet the 1500 ppm threshold set by HCSP |
|---|---|---|---|---|---|---|---|---|
| G | f | 1 | 59 | 23 | 120 | 870 | 891 | 67 |
| G | f | 2 | 59 | 24 | 120 | 861 | 886 | 72 |
| G | b | 1 | 44 | 24 | 120 | 1980 | 2353 | 18 |
| F | a | 1 | 24 | 16 | 90 | 2711 | 2935 | 8 |
| F | a | 2 | 24 | 16 | 90 | 2686 | 2848 | 8 |
| G | a | 1 | 41 | 22 | 120 | 2107 | 2298 | 15 |
| I | a | 1 | 17 | 20 | 240 | 1722 | 1828 | 18 |
| B | a | 1 | 45 | 13 | 90 | 2119 | 2290 | 9 |
| G | d | 1 | 42 | 32 | 110 | 2228 | 2375 | 20 |
| E | a | 1 | 31 | 12 | 120 | 2422 | 2729 | 7 |
| G | e | 1 | 70 | 32 | 120 | 1533 | 1804 | 34 |
| H | a | 1 | 30 | 24 | 180 | 2121 | 2849 | 16 |
| C | a | 1 | 35 | 13 | 80 | 2461 | 2789 | 7 |
| G | c | 1 | 37 | 14 | 240 | 1471 | 1906 | 16 |
| A | a | 1 | 48 | 22 | 180 | 3904 | 4393 | 7 |
| J | a | 1 | 22 | 22 | 180 | 1275 | 2659 | 31 |
| D | a | 1 | 40 | 21 | 100 | 2737 | 3343 | 10 |
| D | a | 2 | 40 | 21 | 100 | 3988 | 4188 | 7 |

| | | | | | | | | |
|---|---|---|---|---|---|---|---|---|
| D | a | 3 | 40 | 20 | 90 | 2661 | 2661 | 10 |
| D | a | 4 | 40 | 18 | 90 | 3069 | 3675 | 8 |

**Table SM.1.** Summary of measurements in the 15 rooms without mechanical ventilation.
