## Supplementary figures and images for "Indoor air quality at a French university: *a participatory CO2 measurement campaign highlights the wide gap between reality and the law*"

### Figure SM.1 Photos and dimensions of room Xa, a room without mechanical ventilation where concentrations above 5,000 ppm were frequently recorded

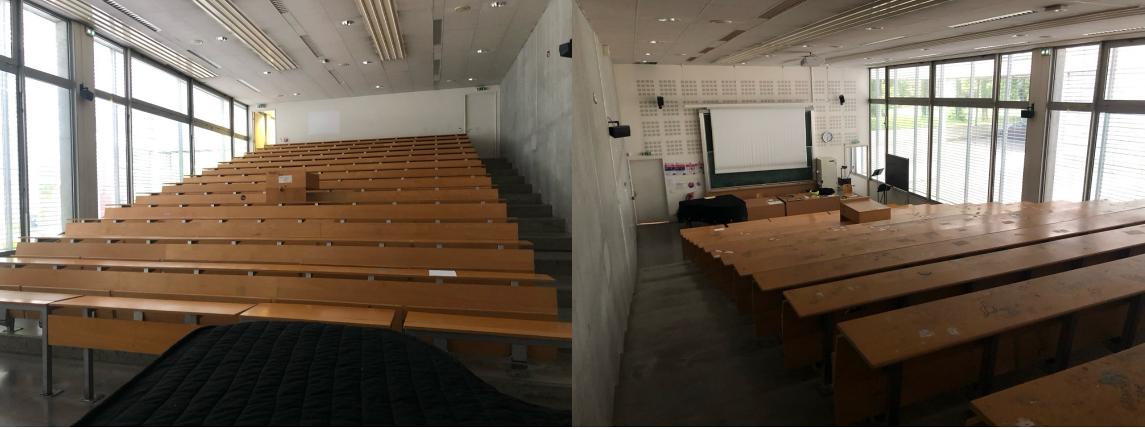

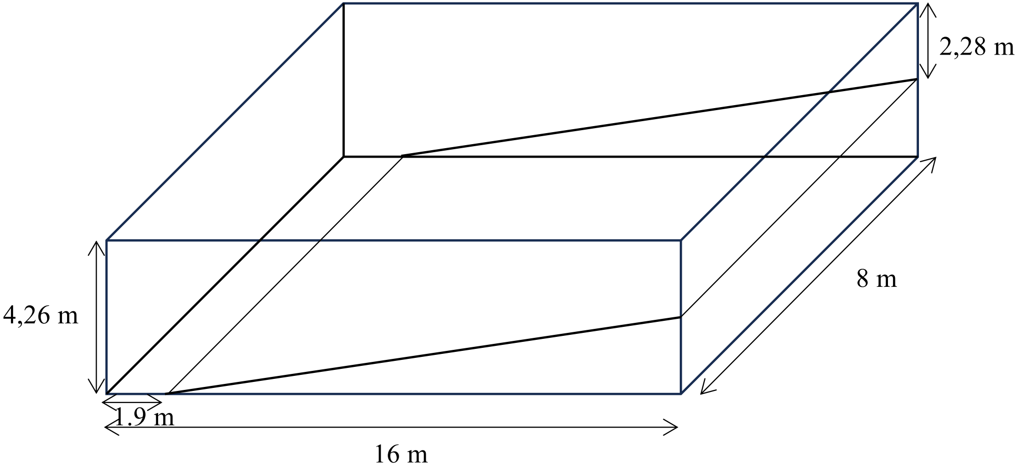

### Figure SM.2 Photos and dimensions of room Vc, where the air handling unit malfunctioned, resulting in CO2 concentrations above 10,000 ppm

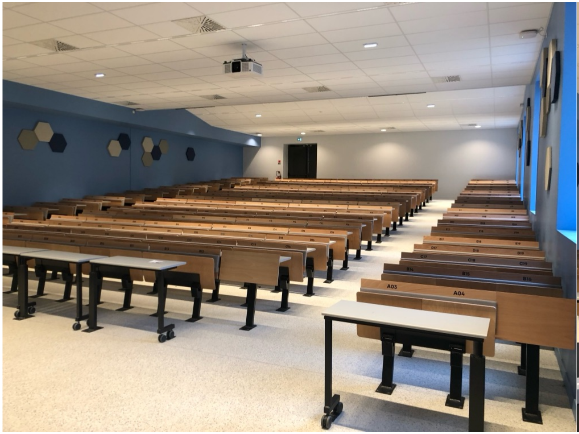

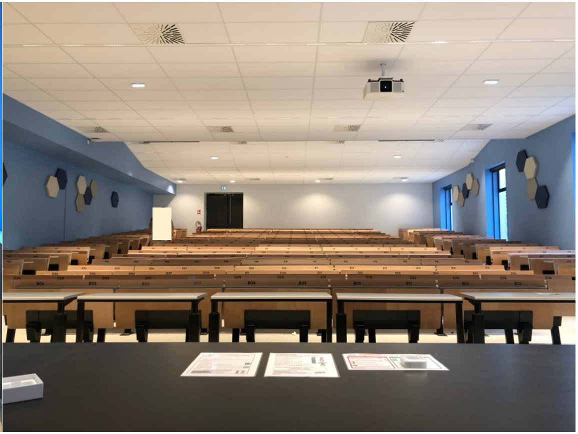

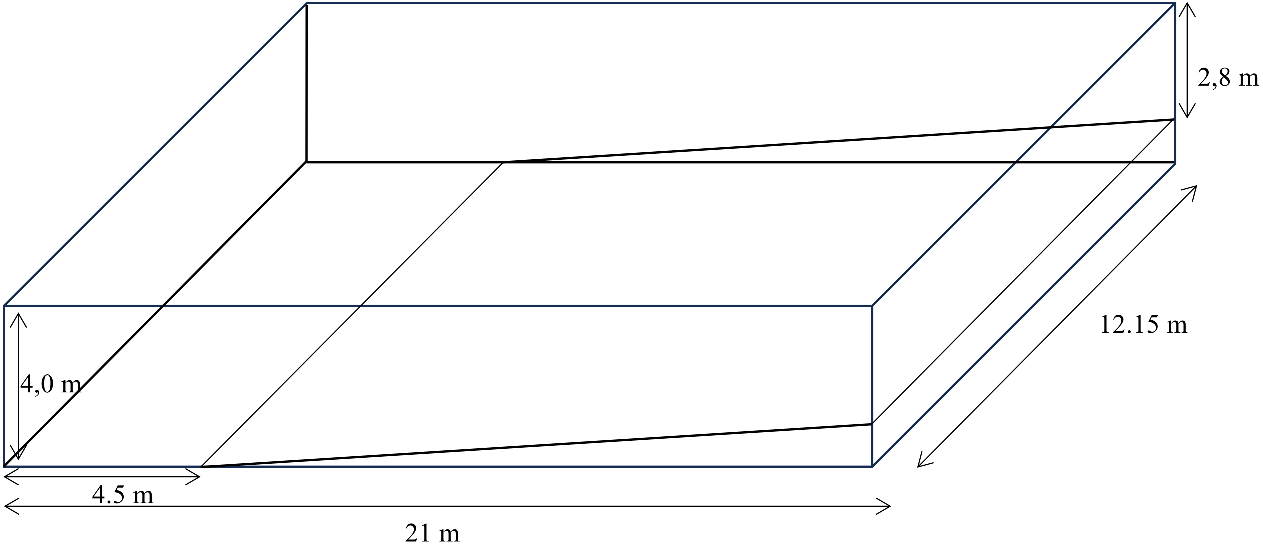
